# Unraveling varying spatiotemporal patterns of dengue and associated exposure-response relationships with environmental variables in Southeast Asian countries before and during COVID-19

**DOI:** 10.1101/2024.03.25.24304825

**Authors:** Wei Luo, Zhihao Liu, Yiding Ran, Mengqi Li, Yuxuan Zhou, Weitao Hou, Shengjie Lai, Sabrina L Li, Ling Yin

## Abstract

The enforcement of COVID-19 interventions by diverse governmental bodies, coupled with the indirect impact of COVID-19 on short-term environmental changes (e.g. plant shutdowns lead to lower greenhouse gas emissions), influences the dengue vector. This provides a unique opportunity to investigate the impact of COVID-19 on dengue transmission and generate insights to guide more targeted prevention measures. We aim to compare dengue transmission patterns and the exposure-response relationship of environmental variables and dengue incidence in the pre- and during-COVID-19 to identify variations and assess the impact of COVID-19 on dengue transmission. We initially visualized the overall trend of dengue transmission from 2012-2022, then conducted two quantitative analyses to compare dengue transmission pre-COVID-19 (2017-2019) and during-COVID-19 (2020-2022). These analyses included time series analysis to assess dengue seasonality, and a Distributed Lag Non-linear Model (DLNM) to quantify the exposure-response relationship between environmental variables and dengue incidence. We observed that all subregions in Thailand exhibited remarkable synchrony with a similar annual trend except 2021. Cyclic and seasonal patterns of dengue remained consistent pre- and during-COVID-19. Monthly dengue incidence in three countries varied significantly. Singapore witnessed a notable surge during-COVID-19, particularly from May to August, with cases multiplying several times compared to pre-COVID-19, while seasonality of Malaysia weakened. Exposure-response relationships of dengue and environmental variables show varying degrees of change, notably in Northern Thailand, where the peak relative risk for the maximum temperature-dengue relationship rose from about 3 to 17, and the max RR of overall cumulative association 0-3 months of relative humidity increased from around 5 to 55. Our study is the first to compare dengue transmission patterns and their relationship with environmental variables before and during COVID-19, showing that COVID-19 has affected dengue transmission at both the national and regional level, and has altered the exposure-response relationship between dengue and the environment.

**Author Summary:** Dengue fever is a typical tropical disease transmitted via mosquito bites. COVID-19 lockdowns have altered human-mosquito contact patterns that impacted dengue transmission. Additionally, lockdowns caused short-term environmental changes that affected dengue vector breeding. In fact, during the COVID-19 period, the normal prevention and treatment of dengue in many dengue-endemic countries was negatively affected due to the sweep of COVID-19, such as strained allocation of medical resources and misreporting of cases. Therefore, this offers a unique chance to study the impact of COVID-19 on dengue transmission, guiding targeted and reasonable prevention measures. We used a series of analytical approaches including time series analysis, space-time scan statistics, and distributed lag non-linear model to compare the differences in dengue transmission patterns and its exposure-response relationships with four environmental variables (average monthly precipitation, average monthly relative humidity, monthly maximum temperature, and monthly minimum temperature) before and during COVID-19 in three Southeast Asian countries: Malaysia, Singapore and Thailand at the province scale. We found that the dengue transmission pattern and its relationship with the environmental variables changed differently. For instance, seasonality and infections heightened in Singapore during COVID-19 and peak relative risk between max temperature and dengue has rose significantly in Northern Thailand.

## 1. Introduction

Dengue, which is caused by four serotypes of the virus (DENV 1-4) and transmitted by mosquitoes, produces flu-like symptoms such as fever and headache (World Health Organization, 2014). The ecology of dengue is shaped by environmental factors that impact the dynamics of vectors, the development of the agent, and interactions between mosquitoes and humans (Morin et al., 2013). It has become a growing threat due to the increasing number of reported cases and deaths worldwide. The World Health Organization (WHO) reports that cases rose from 505,430 in 2000 to 5.2 million in 2019, and deaths more than quadrupled from 960 in 2000 to 4,032 in 2015 (World Health Organization, 2014), significantly burdening public health and socio-economic development in many countries (Srisawat et al., 2022). The onset of the COVID-19 pandemic in 2020 brought about significant societal disruptions and complicated the transmission of dengue (Chen et al., 2022; Wilder-Smith, 2021). To curb the spread of the virus, countries introduced diverse public health measures such as quarantine, isolation, social distancing, and mask-wearing (Lai et al., 2020; Yin et al., 2021). These interventions, coupled with the public’s fear of contracting COVID-19, prompted many individuals to remain indoors, thereby altering human mobility and contact patterns (Xiong et al., 2020; Zhu et al., 2022). Since close proximity contact among human-mosquito contact is a vital factor in dengue transmission (Castro et al., 2021; Salje et al., 2021; Wesolowski et al., 2015; Zhu et al., 2019), but it is decreased in 2020 due to the COVID-19-related interventions, such as in Americas and SEA (Chen et al., 2022; World Health Organization, 2014). However, many scholars believe otherwise that the pandemic actually deteriorated dengue transmission as it strained healthcare systems and limited resources for dengue prevention and vector control (Chen et al., 2022; Olive et al., 2020; Reegan et al., 2020). The declining trend shown by the data is merely the result of under-reporting of dengue cases during the pandemic (Olive et al., 2020).

SEA is an endemic region for dengue (Colón-González et al., 2023; Ooi and Gubler, 2009; van Panhuis et al., 2015) approximately with 2.9 million dengue episodes and 5,906 dengue-related deaths occurring annually. The region suffers from an economic burden of roughly $950 million from dengue (Shepard et al., 2013). During the COVID-19 pandemic, the number of dengue cases in the region decreased, but some countries like Singapore experienced an increase in cases (Chen et al., 2022; Huang et al., 2022). These inconsistent patterns at the regional and national levels indicate a complex situation that requires further investigation.

Researchers are paying attention to the complex impact of COVID-19 and related interventions on dengue transmission in SEA to which both COVID-19 and dengue pose a heavy burden. Ong and Mohd (Ong et al., 2021) utilized a Seasonal Autoregressive Integrated Moving Average (SARIMA) model to investigate the implications of lockdown on dengue transmission in Malaysia. They discovered a possible diffusive effect of dengue vector, which accelerated the increase in dengue incidences. Additionally, Chen et al. (Chen et al., 2022) quantified the effects of COVID-19-related disruptions on dengue transmission in SEA and Latin America, revealing a strong relationship between interventions and reduced dengue risk. Furthermore, the COVID-19 interventions has indirectly led to short-term environmental change (Yip et al., 2022). Thus, the relationship between dengue incidence and environmental variables are susceptible to indirect effects of COVID-19. The exposure-response relationship between environmental variables and the dengue incidence has received widespread attention (Aswi et al., 2018). Zhiwei Xu et al. (Xu et al., 2019) conducted a zonal investigation into the lagged effects of mean temperature and relative humidity on dengue incidence in Thailand by using DLNM from 1999 to 2014, revealing significant relationship between the two factors but noteworthy spatial differences.

Due to the recent emergence of the pandemic, a decrease in government attention to dengue or instances of misjudgment and under-reporting may result in limited availability of accurate dengue cases data(Lam et al., 2020; Yan et al., 2020). Many of the existing studies only focus on one country or province (Li et al., 2021; Lim et al., 2020; Liyanage et al., 2021; Ong et al., 2021; Saita et al., 2022). Although a few studies have attempted to approach the question at a regional level, but they overlooked the analysis at a finer spatial scale (e.g. first-level administrative), as observed in Chen et al. (Chen et al., 2022). This limitation neglects potential regional synchrony and spatial heterogeneity, overestimates the significance of observations, and impedes the generation of insights that can be applied to a large area with a finer spatial scale. Furthermore, the majority of current studies concentrate solely on dengue incidences during the pandemic as a means of comparison to investigate the impact of COVID-19 (Huang et al., 2022; Surendran et al., 2022).

However, dengue incidences can be cyclical (Saita et al., 2022) so restricting the study period solely to the pandemic duration has the potential to introduce bias into the results. As highlighted by Chen et al. (Chen et al., 2022), 2019 experienced the most significant global outbreak of dengue in history, which can complicate the observation and attribution of the effects of COVID-19 disruption. Dengue transmission varies annually due to both seasonal and cyclic patterns. Given the impact of the pandemic on dengue transmission during the past three years, it is imperative to consider all three years comprehensively. However, most existing studies have focused solely on either 2020 or 2021 without taking a holistic approach. In addition, existing research predominantly employs quantitative analysis methods to investigate the influence of environmental variables on dengue, yet overlooks the indirect impacts of COVID-19 lockdown measures on short-term environmental change may result in shifts in the relationship between environmental variables and dengue during the pandemic (Bonnin et al., 2022; Cheng et al., 2021; Chien and Yu, 2014; Chuang et al., 2017; Gui et al., 2021; Sarma et al., 2022; Wang et al., 2022; Xu et al., 2019; Yip et al., 2022).

Therefore, to gain a complete understanding of the impact of the pandemic on dengue transmission, this study aims to address knowledge gaps regarding the impact of COVID-19 and related disruptions on dengue fever transmission and the exposure-response relationship between environmental variables and dengue cases in SEA, with a specific focus on Thailand, Malaysia, and Singapore. To achieve this, we categorized dengue cases and related environmental data into pre-COVID-19 (January 2017 to December 2019) and during-COVID-19 (January 2020 to December 2022). By doing so, we gained a comprehensive comparison and understanding of the two periods of the spatio-temporal distribution of dengue cases in the region and explored the pandemic’s role in its spatial variations using time-series analysis. DLNM was then used to analyze the different relationship of environmental variables with dengue incidences in the two periods taking the impact of lockdown measures on human mobility into consideration. Data from 2012-2017 was also used to visualize and analyze overall trends in dengue cases.

Our study provides new insights into how dengue outbreaks have evolved in these three SEA countries over one decade from 2012 to 2022 and the impact of COVID-19 on dengue to generate a deeper understanding of dengue infection. By informing policies that improve the efficiency of preventing and monitoring dengue cases in the during-COVID recovery era, we hope to contribute to the fight against this disease.

## 2. Methods

### 2.1. Data

Our study focuses on the dengue situation in SEA. Despite countries’ efforts to monitor dengue cases, not all data is readily available for public access. Due to limited data availability, we collected dengue surveillance data and environmental data for three countries at the provincial level from 2012 to 2022, including Singapore, Malaysia, and Thailand, out of 11 SEA countries. With the exception of Singapore, which only provides data at the country level, provinces represent the first administrative tier. As Singapore’s population and land areas are similar to some provinces in other countries, Singapore’s country-level data can be compared with Malaysia’s and Thailand’s province-level data (Singapore department of statistics, 2023; The official portal of sarawak government, 2023).

Dengue data for Malaysia were provided by the Malaysia Ministry of Health (2010 - 2022). Dengue data for Thailand were provided by the Thailand Bureau of Epidemiology (2003 - 2022) and the data for Singapore were provided by the Singapore Ministry of Health (2012 - 2022). For both Thailand and Singapore, weekly dengue data is available whereas for Malaysia, only monthly data is provided. For standardization, dengue data for both Thailand and Singapore are aggregated to obtain monthly data. In addition, it is noteworthy that data spanning from August 2015 to May 2016 in Malaysia were absent in the dataset. To address this gap, we supplemented the dataset utilizing data from a previous study conducted by Chen Y et al (Chen et al., 2022). Moreover, employing Google Earth Engine, we gathered environmental variables in the study region spanning from 2017 to 2022. We selected four environmental variables that have been confirmed to be associated with dengue transmission (Morin et al., 2013; Naish et al., 2014; Xu et al., 2016), including average monthly precipitation, average monthly relative humidity, monthly maximum temperature, and monthly minimum temperature. We also collected Non-Pharmaceutical Interventions (NPI) data (Oxford COVID-19 Government Response Tracker (OxCGRT) project’s overall stringency index) from 2020 to 2022 that serves as a fixed variable in environmental-related analyses, facilitating the simulation of more authentic human mobility patterns during the COVID-19 period. This index, derived from various restrictions related to public health and social measures, and the value ranges from 0 to 100, with higher values representing stronger lockdown measures.(Hale et al., 2021)

### 2.2. Time Series Analysis

We perform temporal variation analysis on the dengue fever data. For each country, the line graph and heatmap are used for better visualization of an overall trend during the study period, and box plots are used for more precise visualization of variations in yearly trends and seasonality. Due to the high seasonality nature of dengue fever (Hartley et al., 2002), we apply seasonal and trend decomposition using loess (STL) to decompose the time series of each country. STL separates time series data into three components: trend (representing long-term and low-frequency variations), seasonal (capturing variations within the same period), and random or remainder (accounting for residual variations after extracting the trend and seasonal components) (Cleveland et al., 1990). Its advantages lie in its simplicity, robustness of results, and effectiveness in data visualization. The equation can be described as follows.

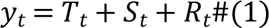

In this context, yt represents the response variable (i.e., case counts) at time t, while Tt, St, and Rt correspond to the trend, seasonal, and remainder components at the same time point. In addition, we also conducted retrospective space-time scan statistics to delineate the spatiotemporal patterns of dengue transmission before and during the COVID-19 (see supplementary appendix for more details).

### 2.3. Exposure-response relationship between environmental variables and dengue fever incidence

DLNM offers robust methodologies for exploring the exposure-response relationship between variables from distinct domains. Particularly within the realm of environmental variables, its reliability surpasses that of traditional time series analyses (Gasparrini, 2011; Gasparrini et al., 2017). DLNM proves to be an invaluable tool for investigating the lagged relationship of dengue fever incidence with environmental variables (Cheng et al., 2021; Chien and Yu, 2014; Chuang et al., 2017; Gui et al., 2021; Sarma et al., 2022; Wang et al., 2022; Xu et al., 2019; Yip et al., 2022). Leveraging the ‘dlnm’ package in R Studio, we conducted comprehensive analyses to compare exposure-response relationship between environmental variables and dengue fever incidence both pre-COVID and during-COVID periods. This approach aims to contribute to a deeper understanding of the impact of environmental variables on the dengue fever incidence.

In this study, the analysis is divided into two distinct phases. Throughout these phases, we employed four key environmental variables (average monthly precipitation, average monthly relative humidity, monthly maximum temperature, and monthly minimum temperature) to illustrate and elucidate the processes under investigation. In the first phase, individual models were constructed for each subregions, aiming to explore the relationship between environmental variables and dengue incidences. A cross-basis was employed for this purpose, utilizing a B-spline with two degrees of freedom (dfs) for the environmental variable space. The specific spline related to environmental variables was positioned at the point representing the lowest risk of dengue. Given the diminished human mobility due to COVID-19 lockdowns, our analysis of the environment-dengue exposure-response relationship incorporates the OxCGRT project’s overall stringency index as a fixed variable. To accommodate seasonal variations and long-term trends within the model, both month and year were integrated as dummy variables. Moving on to the second phase, a multivariate meta-analysis approach was embraced to synthesize the relationship between environmental variables and dengue incidence across diverse areas. This process involved identifying specific associations, considering various lag times for the regions studied. The initial phase of analysis was governed by the following mathematical model:

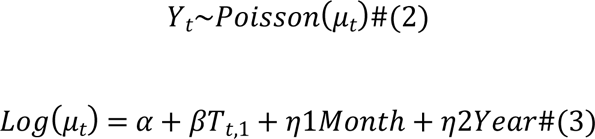

In the provided equation, where t represents the month, Yt signifies the number of dengue cases in a given month, α denotes the model intercept, and β is the vector of coefficients for Tt,1 and the lag month. Tt,1 represents the matrix derived through DLNM for each type of environmental factor.

For analyzing the exposure-response relationship between dengue incidence and environmental variables, given the substantial geographical expanse of Thailand and the comparatively smaller size of Singapore, we partitioned Thailand into six distinct study areas based on classification out of meteorological purposes by the Thai Meteorological Department (Thai Meteorological Department, 2023), while Malaysia and Singapore constituted a singular study area. The delineation of these study areas is visually depicted in Fig. S1.

## 3. Results

### 3.1. Overall Trends

We found strong cyclic patterns in dengue fever incidences across all three countries in the past decade. Fig. 1 depict the annual trends of dengue cases at the national level from 2012 to 2022. Showing that a year with severe dengue situation was often followed by 1-2 years of relatively fewer cases, indicated by the appearance of spikes every 2-3 years. For Singapore, the low peaks occurred in 2015, 2017, and 2021 with high peaks observed in 2013, 2016, 2020 and 2022. For Malaysia, low peaks took place in 2016, 2018, and 2021 with high peaks in 2014, 2017, and 2019. For Thailand, low peaks were observed in 2014, 2017, and 2021 with high peaks in 2013, 2015, and 2019. Except when dengue cases all dipped in 2021 for the three countries due to COVID-19 related interventions, the cyclic patterns rarely overlapped, highlighting unique temporal patterns within each country.

**Fig. 1.**
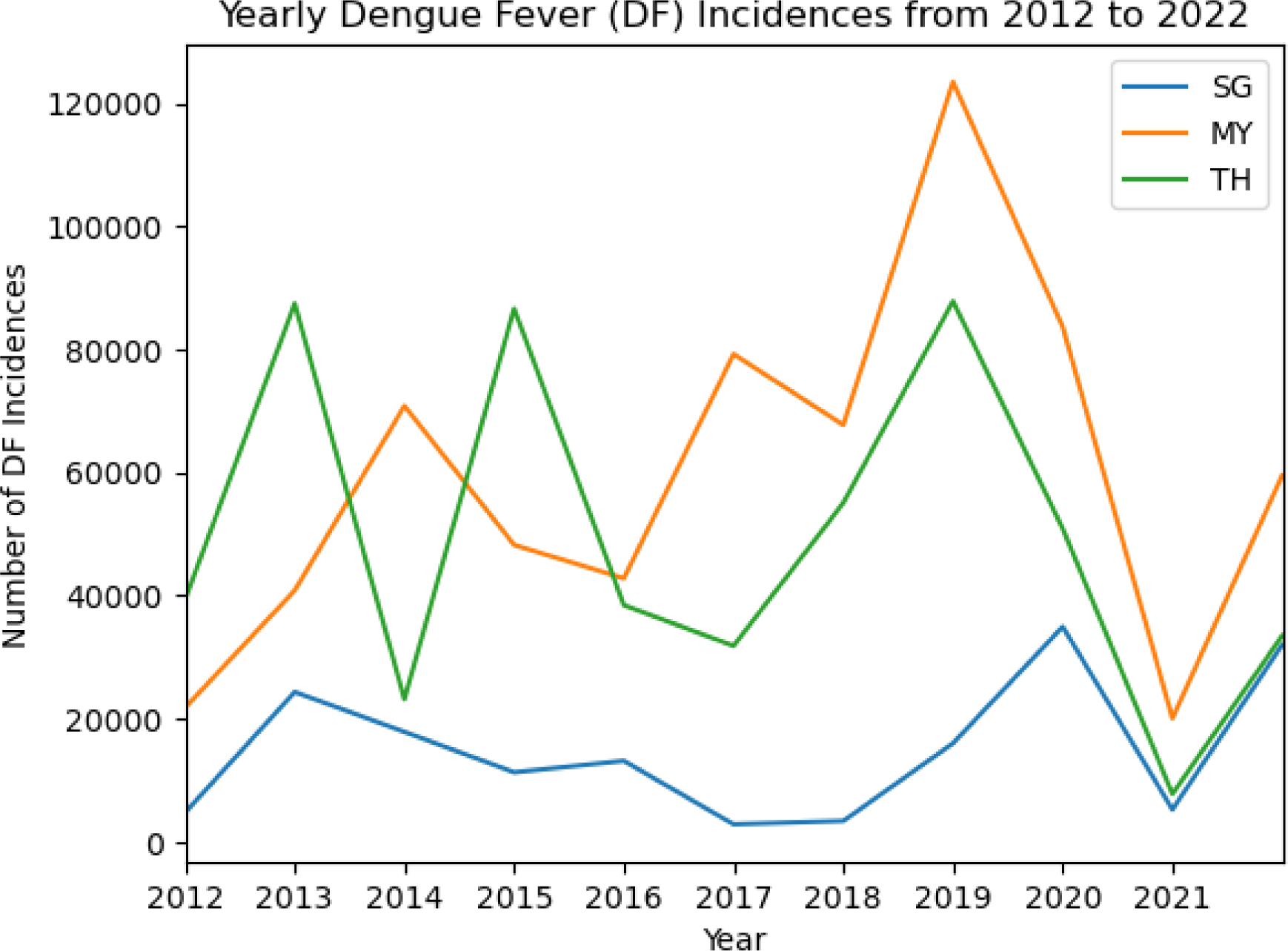
Annual dengue trend in three countries

From a subregional perspective (Fig. 2), all subregions in Thailand exhibited remarkable region-wide synchrony with a similar annual trend except 2021 due to impact from COVID-10-related lockdowns. Furthermore, the pattern of dengue transmission exhibits interannual seasonality. Upon closer observation, the peak incidence of dengue in Northern and Northeastern Thailand occurs earlier and concludes sooner each year compared to other subregions of the country. In relative terms, Malaysia and Singapore have not exhibited pronounced similar trends; however, the variation trend between 2017 and 2022 similar to Thailand.

**Fig. 2.**
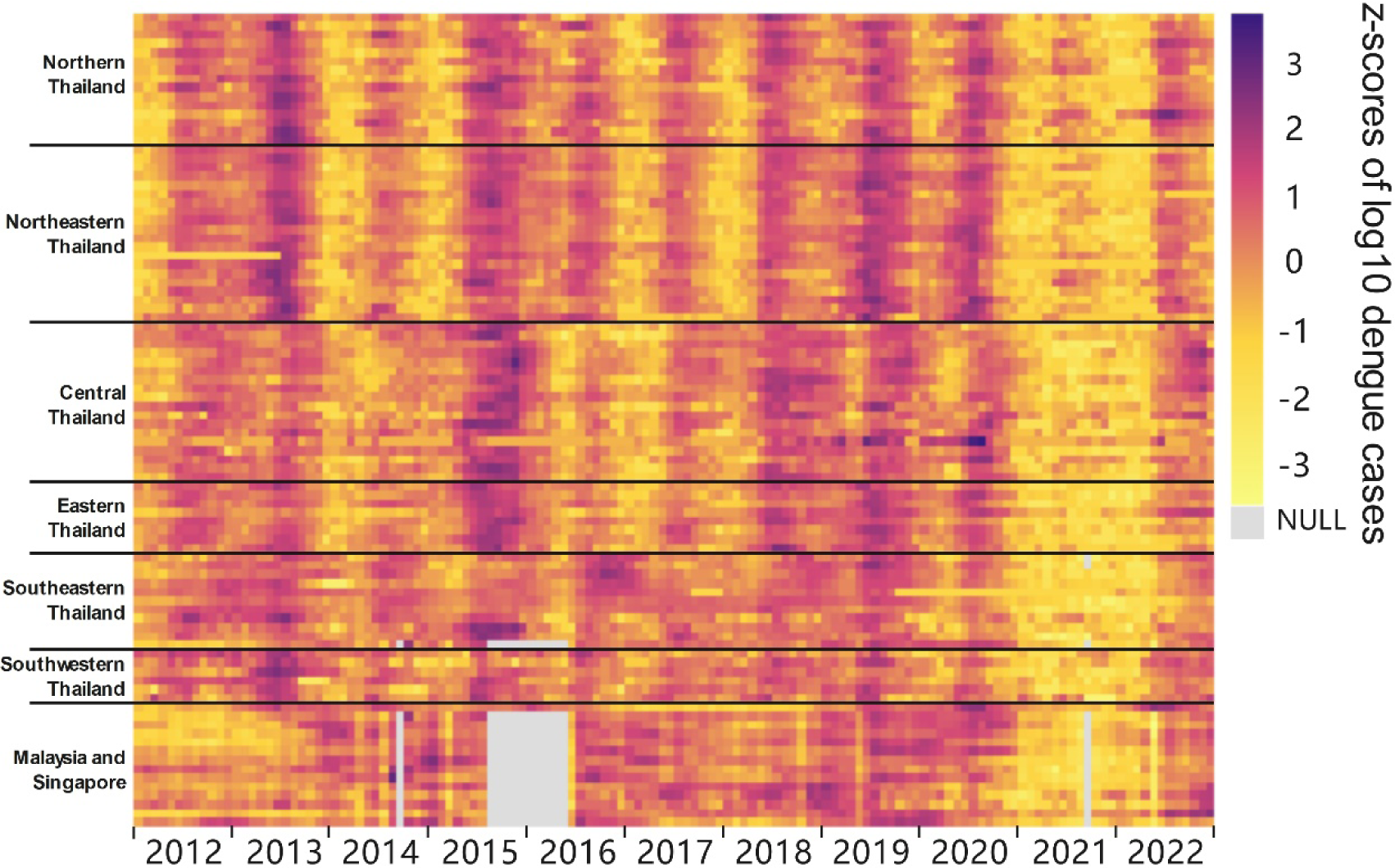
Monthly dengue trend in all subregions.

### 3.2. Time Series Analysis for Seasonal Variations

With STL, we decompose the dengue incidences into yearly trends, seasonal variations, and residue shown in Fig. 3. The cyclic patterns observed in the trend components coincide with those shown in Fig.2 whereas the seasonal components highlight the seasonality of dengue cases, which often rise during the hot and rainy seasons of a year (Colón-González et al., 2013).

**Fig. 3.**
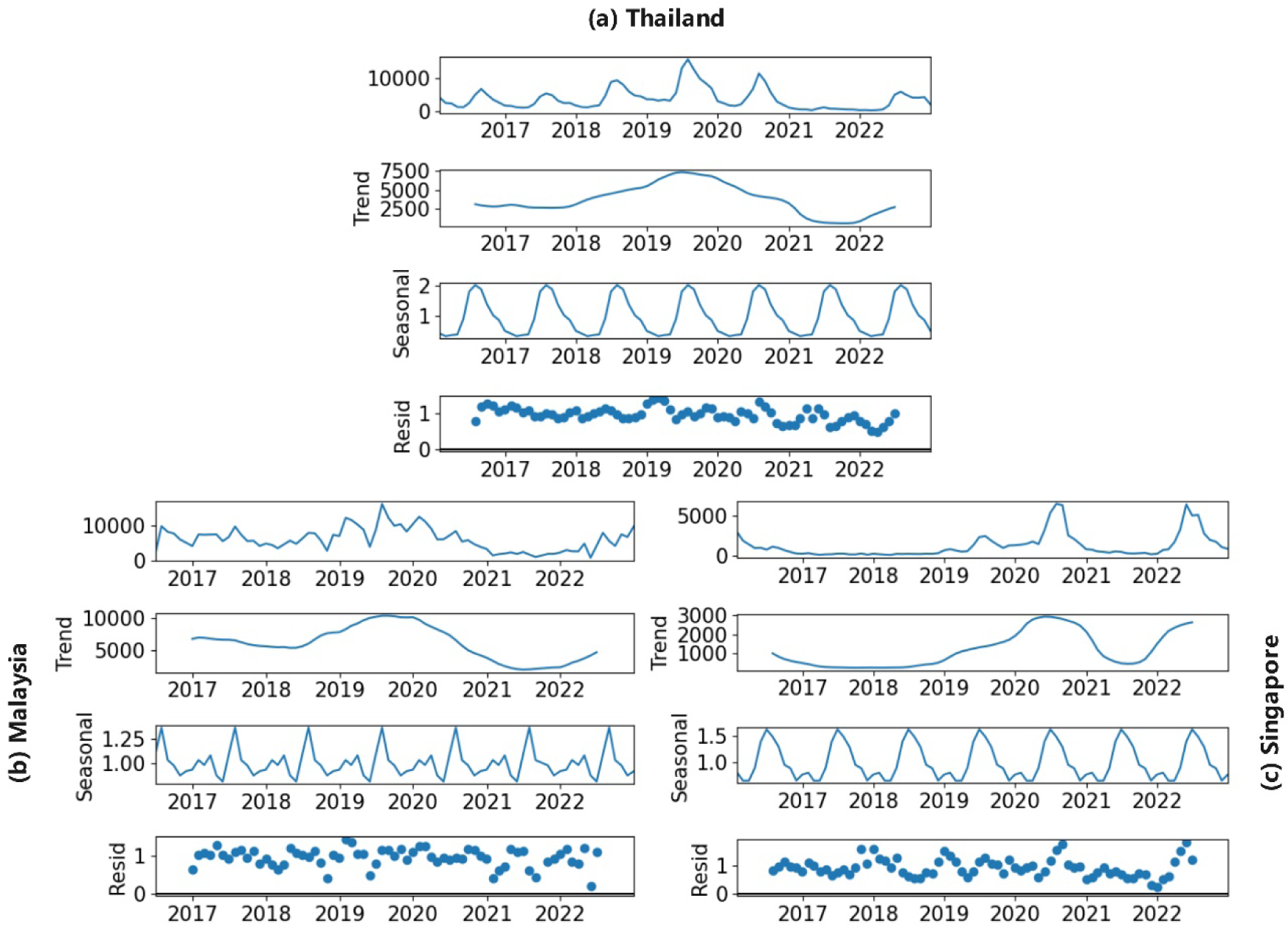
Seasonal Patterns of Dengue Fever Incidences

To better capture seasonality and compare the impact of COVID-19, we use box plots to depict variations in each month’s dengue cases, shown in Fig. 4. Interesting patterns emerge across the three countries by comparing the seasonality plots before (2017-2019, shown by the left column) and during (2020-2022, shown by the right column) COVID-19. For Singapore, dengue cases usually spiked sharply between May and August, which is consistent before and during the pandemic. However, the seasonality impact intensified with the presence of COVID-19: average monthly cases in the middle of a year increased from around 1,250 to 5,000. A high-risk cluster (RR=13.81) was also detected from April 2022 to October 2022 in Singapore (Fig. S7 & Table. S2). For Malaysia, a general reduction in dengue cases throughout the year can be witnessed, coupled with heightened variability between same months during the COVID-19 period. Meanwhile, the low-risk cluster in Western Malaysia expanded and witnessed a reduction in RR (from 0.34 to 0.10) (Fig. S7 & Table. S1-S2). For Thailand, average monthly cases during dengue season, i.e., the middle of a year, showed slight decrease. A low-risk cluster (RR=0.08) was identified in Southern Thailand accordingly, while a high-risk cluster (RR=71.07) emerged in Mae Hong Son during dengue season (Fig. S7 & Table. S2). This was probably due to Mae Hong Son implemented sheltered sanitation practices and experienced the maximum temperature and miserable humidity levels, promoting the mosquito breeding (Saita et al., 2022).

**Fig. 4.**
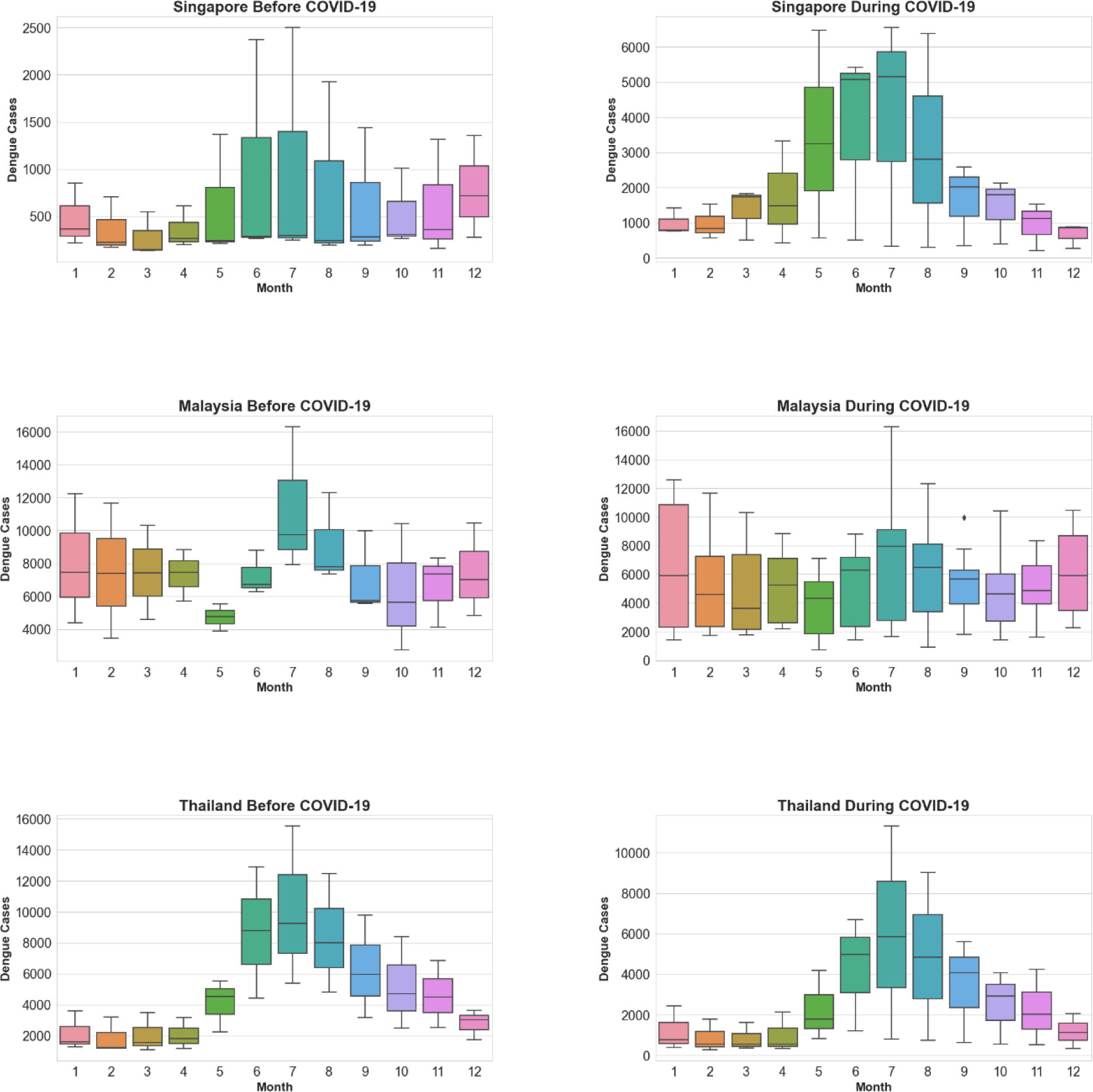
Box Plots of Seasonal Patterns of Dengue Fever Incidences Before and After COVID-19

### 3.3. Exposure-response relationship between environmental variables and dengue fever incidence in pre-COVID-19 and during-COVID-19

We employ DLNM to unveil the exposure-response relationship between various environmental variables and dengue fever, both pre-COVID-19 and during-COVID-19, across different study areas. Meanwhile, the overall cumulative association with a lag of 0-3 months was explored between environmental variables and dengue fever cases in the same contexts.

In a comprehensive overview, the overall exposure-response relationship between dengue cases and environmental variables in total region of Thailand, as well as Malaysia and Sinagpore exhibited different changes (Fig. 5). For Thailand, during the pandemic, the 0-1.5 months following the attainment of maximum temperatures between 25℃-30℃ emerge as new occurrences of high-risk periods. In contrast, before pandemic, the RR within this maximum temperature range was exceedingly low. For Malaysia and Singapore in pre-COVID-19, the high RR interval for relative humidity was 1.5-3 months after reaching 80%-90%, with the peak value at 2.7. There was a significant shift in this pattern during pandemic, with the high RR lag interval occurring approximately 0.4-3 months after relative humidity reached 80%-90%. This suggests a rapid increase in RR when relative humidity reaches this interval, with an earlier and sustained lag period and a significantly heightened sensitivity. However, the highest value of RR significantly decreased, dropping from 2.7 to 1.4. For minimum temperature, the high RR interval was relatively broad before pandemic, occurring 0-3 months after the temperature reached around 20℃-23℃. It is noteworthy that this lagged relationship differs from the majority, as the RR sharply increases when the minimum temperature enters this interval, gradually decreasing over time, contrary to the typical pattern where RR tends to gradually rise with increasing lag duration. During COVID-19, the high-risk range has narrowed significantly and the RR has also decreased. Moreover, the overall cumulative association between environmental variables and RR in Thailand exhibited substantial changes with a lag of 0-3 months. Specifically, the peak of the overall cumulative association for precipitation and dengue cases decreased by approximately 0.3. The association between relative humidity and dengue cases underwent a significant transformation. Pre-pandemic, the curve exhibited a slightly inverted U-shape, peaking at around 80%, with a value of approximately 1.0. However, during the COVID-19, the associations between relative humidity and RR became proportional, steadily increasing above around 80%, instead of decreasing. The maximum temperature curve has transitioned from an inverted U-shape to a positive correlation as temperatures reach their peak. For minimum temperature, in comparison to the pre-pandemic period, a conspicuous decrease in RR has been observed when the minimum temperature exceeds 23 ℃. For Malaysia and Singapore, the characteristics of the association between dengue and precipitation, relative humidity, exhibit interestingly reverse patterns in the perspectives of two periods compared with Thailand. While the association with the maximum temperature exhibits marginal changes, the RR of the association between dengue and the minimum temperature, specifically within the range of 20℃-22℃, shows a relatively distinct decrease.

**Fig. 5.**
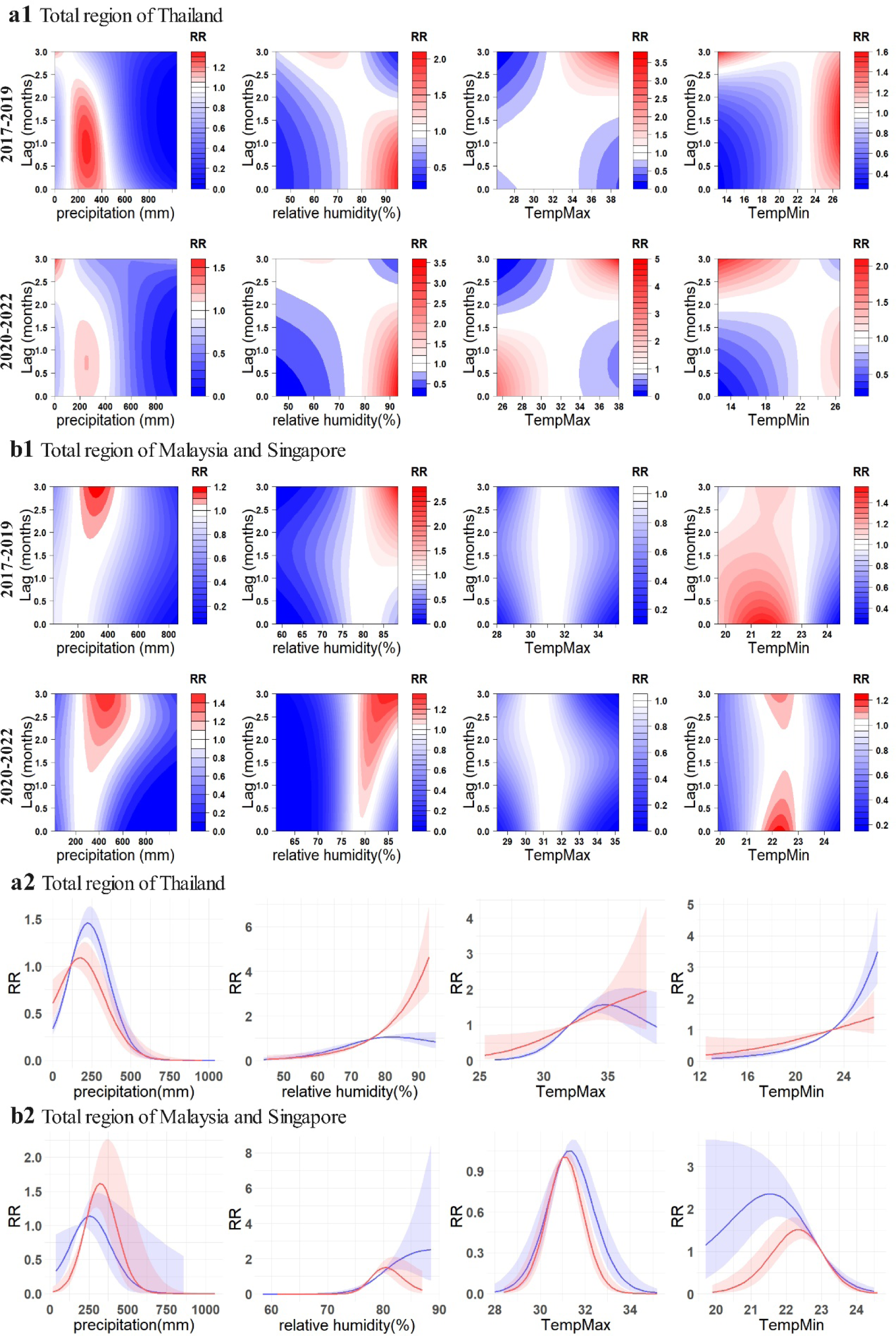
Exposure-response relationship between dengue incidences and environmental variables in total region of research area. For a1 and b1, contour plots showcase the relationship between four environmental variables and dengue RR in Thailand, Malaysia, and Singapore respectively. a2 and b2 illustrate the overall cumulative associations (0-3 months) between the four environmental variables and dengue RR in Thailand, Malaysia and Singapore. The blue lines represent the pre-COVID-19 (2017-2019), while the red lines represent the during-COVID-19 (2020-2022), with the corresponding 95% confidence intervals indicated in similar colors.

At the subregional scale in Thailand, the environmental variables with the exposure-response relationship to dengue fever that varied considerably before and during the pandemic include precipitation, maximum temperature and minimum temperature in Northern Thailand, relative humidity in Southeastern Thailand, minimum temperature in Northeastern Thailand and Southeastern Thailand, and precipitation in Southwestern Thailand.

By contrast, the exposure-response relationship of environmental variables with dengue cases were most variable in Northern Thailand (Fig.6). Specifically, high RR cluster of precipitation shows minimal variations in high RR areas. However, there is a significant elevation in RR values, with the highest value increasing from 2.2 to 7.9. The maximum temperature exhibited the most noticeable change among all environmental variables and regions. Pre-pandemic, the high RR cluster was 0-1 month and 2.5-3 months after temperatures between 27℃-32℃. However, during the pandemic, there was a significant increase in RR within 0-1 month after this temperature range. Additionally, a new high RR zone emerged, 2.5-3 months after temperatures between 34℃-38℃. This suggests that the dengue cases in Northern Thailand were highly influenced by the maximum temperature in these two temperature intervals during the pandemic. Interestingly, the previously identified high RR zone, 2.5-3 months after the pandemic, rapidly decreased to no risk, i.e., 0. For minimum temperature in the same region, the high RR zone also completely shifted its position. Pre-pandemic, the high RR zone was 0-1 month after temperatures between 13℃-21℃. During the pandemic, a new high RR zone emerged, 2.5-3 months after temperatures between 17-22. The 0-3 months cumulative RR association between relative humidity and dengue cases in Northern Thailand changed from a relatively flat curve to a clear U-shape, with a significant increase in RR.

**Fig. 6.**
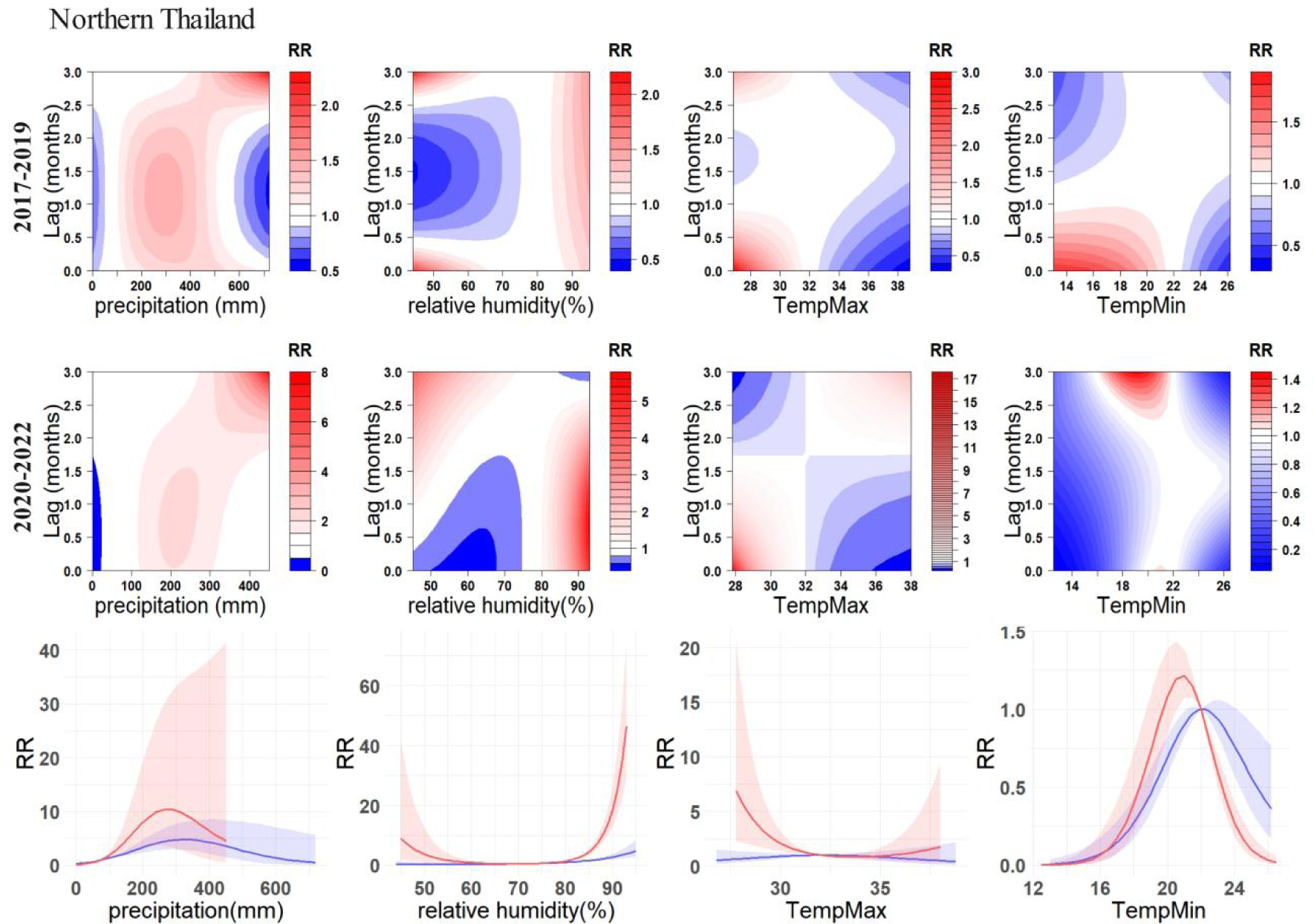
Exposure-response relationship between dengue incidences and environmental variables in Northern Thailand. Contour plots showcase the relationship between four environmental variables and dengue RR. Line charts illustrate the overall cumulative associations (0-3 months) between the four environmental variables and dengue RR. The blue lines represent the pre-COVID-19 (2017-2019), while the red lines represent the during-COVID-19 (2020-2022), with the corresponding 95% confidence intervals indicated in similar colors.

Southeastern Thailand’s relative humidity lagged impact pattern was intricate (Fig. S2). The RR was generally low for relative humidity between 77%-85% during pre-pandemic, whereas the RR significantly increased in the lag intervals of 0-0.3 and 2.5-3 months after reaching this relative humidity range during the pandemic. In addition, 1.8-3 months after the relative humidity range reached 55%-75% before the pandemic was a high RR zone, and during the pandemic, the zone shrank significantly to 2.5-3 months after the relative humidity range reached 60%-70%. Additionally, the previously identified high RR cluster, 2-3 months after relative humidity between 55%-75%, substantially narrowed down to 2.5-3 months after reaching 60%-70%. Notably, the lagged relationship between relative humidity and the RR of dengue cases in Southeastern Thailand exhibited significant variations, forming a marked contrast to the overall trend observed in Thailand. Furthermore, Northeastern Thailand’s minimum temperature high RR cluster completely shifted its position, demonstrating a highly significant change (Fig. S3). Pre-pandemic, the high RR interval was 0-2 months after the temperature reached 16℃-22℃, with the peak RR occurring one month after the lag. However, during the pandemic, this interval transitioned to a low-risk zone, and two new high RR zones emerged: 0-0.5 months and 2.5-3 months after the temperature reached 22℃-26℃. For the Eastern Thailand (Fig. S4), there has been a discernible decline in the RR associated with the maximum temperature in relation to dengue, the maximum value has decreased from approximately 3.5℃ to 1.5℃. Over a cumulative lag of 0-3 months, the peak value for RR of association between precipitation and dengue has risen from approximately 1.8 to 3.4, the precipitation remains relatively constant at its peak. Additionally, the peak value for relative humidity has decreased from about 3.4 to 1.1, accompanied by an approximate 2% increase in relative humidity when the peak occurs. The exposure-response associations between dengue incidences and environmental variables in Central Thailand and Southwestern Thailand changed little before and during COVID-19 (Fig. S5-S6).

## 4. Discussion

### 4.1. Principal Findings

Our study aims to explore the spatio-temporal patterns and exposure-response relationship between environmental variables and dengue fever for both pre-and during-COVID-19. This will be achieved by analyzing the dengue incidence data of three Southeast Asian countries - Thailand, Malaysia, and Singapore in the pre-COVID-19 and during-COVID-19. To the best of our knowledge, this study is the first attempt to utilize province-level data from multiple countries in SEA to generate regional-level insights into dengue fever over the special time period. While previous studies have shown that the situation regarding dengue fever improved during the pandemic at both regional and global levels (Chen et al., 2022; World Health Organization, 2014), the impact can vary across countries. Malaysia and Thailand saw a reduction in the burden of dengue fever during the pandemic, while Singapore experienced unprecedentedly terrible dengue outbreaks in 2020 and 2022 (Ong et al., 2021).

In order to investigate the fluctuations in dengue incidence during the pandemic, we employed time series analysis to examine the seasonality component of the monthly dengue incidence data. We observed that both Singapore and Thailand maintained pronounced seasonality in dengue cases pre-and during-COVID-19. And a notable increase in dengue cases in Singapore during COVID-19, especially from May to August, experiencing an approximately fourfold rise compared to the same period pre-COVID-19. Possibly attributed to increased human-mosquito contact opportunities due to population staying at home, a surge in blood supply (Ong et al., 2021). And the southwestern monsoon (end of May to September) results in elevated temperatures, humidity, and precipitation levels conducive to mosquito proliferation(World Health Organization, 2020). In contrast, Malaysia exhibited a more evenly distributed number of dengue cases across months during COVID-19, indicating a weakened seasonality compared to the before COVID-19. And during COVID-19, both Thailand and Malaysia experienced a relatively significant decline in the dengue cases. Possibly because Malaysia and Thailand potentially implemented divergent lockdown measures during the same period, while the timing and intensity of monsoons may have exhibited variations across the three countries. Furthermore, distinctions in healthcare infrastructure and the differences of public health interventions could contribute to disparate impacts on dengue incidence in Singapore, Malaysia, and Thailand (Malaysia Health Ministry, 2024; Singapore Health Ministry, 2024; Thailand Health Ministry, 2024). Additionally, prior research has observed a slight delay in Thailand’s dengue seasonality (Saita et al., 2022), but our study is the first to focus on the effect of intensified seasonality. While false positive dengue test results in COVID-19 patients have been reported (Lam et al., 2020; Yan et al., 2020), this alone does not completely explain the observed seasonal variations. In 2020, the surge in dengue cases coincided with Singapore’s COVID-19 lockdown period from April to June. However, in 2022, despite the easing of COVID-19 measures, dengue incidences still surged in May. Moreover, as noted by Brady and Wilder-Smith (Brady and Wilder-Smith, 2021), lockdowns are only temporary changes in human mobility and are unlikely to have a lasting impact on vector population or their behavior. Consequently, even in areas that saw a decrease in dengue cases during lockdowns, it is possible that those infections were only postponed rather than prevented. Thus, other factors such as environment, socioeconomic status, and vector distribution should be examined over a more extended study period to establish a causal relationship between stay-at-home orders and dengue surges.

The differences in the lagged relationship between environmental variables and dengue cases before and during the pandemic reveal a more complex set of influencing factors. Previous research has indicated that an increase in temperature promotes the proliferation of Aedes aegypti and Aedes albopictus mosquitoes, enhancing the severity of dengue infections (Gui et al., 2021). Our study indicates that only the RR of dengue cases in Northern Thailand showed a significant increase during the pandemic due to temperature variable. According to our data, the average monthly maximum temperature in Northern Thailand rose from 31.99 ℃ in 2017-2019 to 32.32℃ in 2020-2022, marking an increase of 0.33 ℃. In contrast, the differences in the monthly average maximum temperature in other parts of Thailand, as well as in Malaysia and Singapore, were extremely small, all below 0.05, with the differences in Southeast and Southwest Thailand, Malaysia, and Singapore even below 0.01. Therefore, the significantly soaring RR of the relationship between maximum temperature and dengue in Northern Thailand may be attributed to the fact that the increase in maximum temperature, promoting breeding of Aedes aegypti and Aedes albopictus mosquitoes, and thus leads to an increase in dengue infection. Additionally, concerning the exposure-response relationship between precipitation and the RR of dengue, there was a significant increase in Northern Thailand during the pandemic. The high RR zone in this region was 2.5-3 months after extreme high rainfall (500mm-700mm) before the pandemic. However, during the pandemic, this rainfall range shifted to around 350mm-400mm, with the lag time for high risk remaining roughly the same, resulting in a significant increase in RR, peaking at 7.9. This finding aligns with the observations by Cheng J et al. (Cheng et al., 2021) in Guangzhou, China, indicating a significant increase in dengue risk 6-13 weeks after extreme high rainfall. This observation is consistent with the perspective on vector, wherein mosquitoes exhibit heightened breeding frequency during the rainy season (Ong et al., 2021). Apart from Northern Thailand, variations in precipitation were observed in other study regions, but the changes in RR were generally small. Therefore, the exposure-response relationship between precipitation and dengue RR exhibits regional differences, necessitating further investigation.

### 4.2. Implications and Recommendations

The COVID-19 pandemic has posed one of the most significant public health challenges. Despite the low likelihood of a similar pandemic in the near future, the insights gained from this unique situation regarding dengue transmission can contribute to our knowledge gap and be applied in future dengue control studies (Chen et al., 2022).

First of all, we want to highlight the importance of vector control for dengue prevention. Our study examines the impact of COVID-19 on dengue transmissions in SEA using province-level dengue incidence data from 2017 to 2022, and reveals the country-specific effects of pandemic-related measures on dengue outbreaks. Although measures such as lockdowns can potentially reduce dengue transmission by limiting mobility and socio-economic activities, it is also likely that residents staying at home increase the risk of mosquito breeding in residential areas (Ong et al., 2021), as seen in Singapore. Therefore, future dengue control efforts should prioritize vector control and the prevention of vector breeding (Buhler et al., 2019; Sulistyawati et al., 2023).

Secondly, we urge relevant authorities to strategically allocate their healthcare resources to ensure proper dengue management even during times of emergency. The allocation of healthcare resources has been put to the test during this public health crisis. Many countries struggled to cope with the sudden pressure on limited resources, resulting in neglect of the dengue situation with discontinued vector control and inadequate dengue testing (Aborode et al., 2022; Harapan et al., 2021; Sulistyawati et al., 2023). Relevant authorities can proactively intervene based on the environmental conditions associated with an elevated dengue infection rate. For instance, in areas experiencing increased precipitation, implementing artificial rainfall reduction measures may be a strategic intervention to reduce mosquito breeding rates (Zhou et al., 2023). Additionally, targeted weed control measures could be employed to minimize breeding zones. Moreover, distributing mosquito repellent and nets (Saita et al., 2022) to residents can help protect them from mosquitoes even if vector control measures are not effective.

Thirdly, we urge SEA countries to strengthen their collaborative efforts in combatting dengue due to dengue transmission mechanisms is complex and country-specific. Localized clusters were prevalent during the pandemic. However, as the world enters the post-COVID recovery era, which sees increasing mobility both within and across country borders, cross-country clusters are increasing. This situation may be exacerbated by the regional synchrony of dengue risk, highlighting the need for regional collaboration in dengue control (Servais, 2009). The SEA countries should work together to prevent, detect, and treat dengue, starting with sharing relevant data with each other (Luo et al., 2023, 2022). The detection of specific dengue serotypes in one country should be made known to other countries as soon as possible so that they can adopt more targeted prevention and detection measures to minimize the risk. Successful control of dengue vectors in one country reduces the risk of cross-border transmission, benefiting the entire region.

Last but not least, this study confirms that during the periods of 2017-2019 (pre-COVID-19) and 2020-2022 (during-COVID-19), the exposure-response relationships between various environmental variables and dengue fever incidence underwent changes across different study regions, exhibiting varying magnitudes. These changes may be attributed to the impact of environmental alterations resulting from COVID-19-related lockdown measures or other natural factors(Li et al., 2022; Liu et al., 2024), leading to variations in dengue transmission sources (Bonnin et al., 2022; Cheng et al., 2021). Therefore, we recommend that health-related departments in each country should pay attention to the increase in dengue infections caused by changes of human mobility pattern or short-term environmental changes. They should take early preventive measures to minimize infections. However, the connection between environmental variables and dengue infections is highly intricate, and the changes in dengue infections are often driven by the combined influence of multiple environmental factors (Morin et al., 2013). Hence, relevant departments or researchers should delve deeper into comprehensive studies to better understand these complex relationships.

### 4.3. Limitations

Despite gaining valuable insights, our results have been limited by various obstacles encountered during the studies on dengue transmission. A significant challenge has been the limited availability of dengue incidence data. While many countries have a nationwide surveillance system for dengue, not all of them make their incidence data publicly available. This has significantly restricted the potential for data-driven studies. Even among the countries that do make their data available, many only provide it at a coarse spatial scale, such as the national level, which limits the possibility of uncovering finer details. Therefore, this study was only able to analyze regional trends in SEA using data from three countries that were available at a suitable spatial scale. Hence, we encourage relevant authorities to publish more finer spatial scale data so that future researchers can generalize the findings are a large scale, which could be significant for dengue surveillance and prevention.

Moreover, with the majority of healthcare resources being directed towards combating COVID-19, there is a valid concern that dengue cases may have been under-reported during the pandemic (Olive et al., 2020), which could explain the decrease in cases seen at a global level. However, a recent statistical analysis by Chen et al. (Chen et al., 2022) suggests that the observed variations in dengue fever during the pandemic are unlikely to be solely due to under-reporting. Despite this reassurance, it is important to conduct further studies in the future to explore the possibility and extent of under-reporting during pandemics to minimize data uncertainty and draw significant conclusions about the impact of pandemics on dengue transmission.

Lastly, the global response to COVID-19 measures has led to a reduction in nitrogen oxide emissions, thereby promoting short-term warming (Forster et al., 2020), which could potentially influence the dengue incidences. However, this study did not confirm that the differences in the exposure-response relationships between environmental variables and dengue incidences before and during the pandemic were definitely influenced by COVID-19. Instead, it is plausible that various environmental factors underwent short-term and slight changes for different reasons during these two periods. Further investigation is required to ascertain what factors are statistically significantly related to the changes of exposure-response relationship between environmental variables and dengue over the two time periods.

## 5. Conclusion

By using time series analysis, we were able to identify the unique impact of COVID-19 on dengue transmission in each of the three countries studied. Although all three experienced localized dengue risk during the pandemic, Singapore was particularly affected by an intensified seasonality effect that led to an unprecedented outbreak of dengue fever in 2020. Additionally, the regional synchrony of dengue underscores the importance of collaboration across borders in the fight against fever dengue. Finally, by employing a distributed lag nonlinear model with NPI, we identified varying degrees of spatial differences in the exposure-response relationships between environmental variables and dengue incidences both pre- and during-COVID-19. Our findings highlight that the changes in human mobility patterns resulting from government interventions against COVID-19, coupled with the indirect impacts on short-term environmental change, can exert varying degrees of dengue transmission.

### CRediT authorship contribution statement

**Wei Luo:** Writing - Review & Editing, Writing - Original Draft, Conceptualization, Supervision, Project administration, Funding acquisition. **Zhihao Liu:** Writing - Original Draft, Writing - Review & Editing, Methodology, Software, Formal analysis, Investigation, Data Curation, Visualization. **Yiding Ran:** Writing - Original Draft, Writing - Review & Editing, Conceptualization, Methodology, Software, Formal analysis, Investigation, Data Curation, Visualization. **Mengqi Li:** Methodology, Software, Formal analysis, Investigation, Data Curation, Visualization. **Yuxuan Zhou:** Conceptualization, Investigation. **Weitao Hou:** Data Curation. **Shengjie Lai:** Writing - Review & Editing, Conceptualization. **Sabrina L Li:** Writing - Review & Editing. **Ling Yin:** Writing - Review & Editing.

## Data Availability

All relevant data are in the GitHub of the GeoSpatialX Lab (https://github.com/GeoSpatialX).

## Acknowledgements

This work was supported in part by National University of Singapore FY2020 START-UP GRANT under WBS A-0003623-00-00 and National Natural Science Foundation of China (No.42271475). SL was supported by the National Institute for Health (MIDAS Mobility R01AI160780) and the Bill & Melinda Gates Foundation (INV-024911). It involves data collection, analysis, and interpretations well as publication fee if applicable. We were not precluded from accessing data in the study, and we accept responsibility to submit for publication.

## Supporting information

S1 File. Study areas in the exposure-response relationship between dengue incidence and environmental variables; Figures of exposure-response relationships of subregions of Thailand; Retrospective Poisson Space-Time Scan Statistic.

